# DMARD disruption, disease flare, and prolonged symptom duration after acute COVID-19 among participants with rheumatic disease: A prospective study

**DOI:** 10.1101/2022.02.08.22270696

**Authors:** Michael Di Iorio, Claire E. Cook, Kathleen M.M. Vanni, Naomi J. Patel, Kristin M. D’Silva, Xiaoqing Fu, Jiaqi Wang, Lauren C. Prisco, Emily Kowalski, Alessandra Zaccardelli, Lily W. Martin, Grace Qian, Tiffany Y-T. Hsu, Zachary S. Wallace, Jeffrey A. Sparks

## Abstract

**Objective:** To describe disease-modifying antirheumatic drug (DMARD) disruption, rheumatic disease flare/activity, and prolonged COVID-19 symptom duration among COVID-19 survivors with systemic autoimmune rheumatic diseases (SARDs).

**Methods:** We surveyed patients with SARDs after confirmed COVID-19 at Mass General Brigham to investigate post-acute sequelae of COVID-19. We obtained data on demographics, clinical characteristics, COVID-19 symptoms/course, and patient-reported measures. We examined baseline predictors of prolonged COVID-19 symptom duration (defined as lasting ≥28 days) using logistic regression.

**Results:** We analyzed surveys from 174 COVID-19 survivors (mean age 52 years, 81% female, 80% White, 50% rheumatoid arthritis) between March 2021 and January 2022. Fifty-one percent of 127 respondents on any DMARD reported a disruption to their regimen after COVID-19 onset. For individual DMARDs, 56-77% had any change, except for hydroxychloroquine (23%) and rituximab (46%). SARD flare after COVID-19 was reported by 41%. Global patient-reported disease activity was worse at the time of survey than before COVID-19 (mean 6.6±2.9 vs. 7.6±2.3, p<0.001). Median time to COVID-19 symptom resolution was 14 days (IQR 9,29). Prolonged symptom duration of ≥28 days occurred in 45%. Hospitalization for COVID-19 (OR 3.54, 95%CI 1.27-9.87) and initial COVID-19 symptom count (OR 1.38 per symptom, 95%CI 1.17-1.63) were associated with prolonged symptom duration. Respondents experiencing prolonged symptom duration had higher RAPID3 scores (p=0.007) and more pain (p<0.001) and fatigue (p=0.03) compared to those without prolonged symptoms.

**Conclusion:** DMARD disruption, SARD flare, and prolonged symptom duration were common in this prospective study of COVID-19 survivors, suggesting substantial impact on SARDs after acute COVID-19.

## INTRODUCTION

People with systemic autoimmune rheumatic diseases (SARDs) may have higher risk for severe outcomes during the acute course of coronavirus disease 2019 (COVID-19)(1-4). However, there have been few investigations about the clinical course of COVID-19 survivors with SARDs following the acute clinical course. Decisions made to modify SARD treatment regimens in the setting of acute COVID-19 and direct effects of SARS-CoV-2 infection may impact disease control of the underlying SARD(5, 6).

After the resolution of the acute phase of COVID-19, a subset of people experience persistent symptoms of COVID-19 and a decline in quality of life for many weeks to months, collectively referred to as post-acute sequelae of COVID-19 (PASC, or colloquially as “long COVID”)(7-9). The continued high rates of transmission, morbidity, and mortality experienced during the global pandemic as new variants emerge emphasize the public health need to better understand the long-term implications of COVID-19(10). Particular attention should be given to participants with SARDs who may be vulnerable to PASC given shared features of acute SARS-CoV-2 infection and SARDS, including systemic inflammation, autoimmunity, hypercoagulability, and fibrosis.

In this prospective, multi-center study, we aimed to describe the impact of COVID-19 on SARD participants after resolution of acute infection, including immunosuppressive medication disruption, rheumatic disease flare/activity, and prolonged duration of COVID-19 symptoms using surveys administered to SARD participants systematically confirmed to have had confirmed COVID-19 and survived.

## METHODS

### Study population

We systematically identified all SARD participants with SARS-CoV-2 confirmed by PCR, nucleocapsid antibody, or antigen testing (from March 1, 2020 to November 3, 2021 for this analysis) at Mass General Brigham (MGB) HealthCare system in the greater Boston, Massachusetts area, as previously described(2, 9, 11-14). This was supplemented by referrals from rheumatologists of participants who were diagnosed outside of the MGB system. Presence of prevalent SARD and SARS-CoV-2 were confirmed by medical record review prior to invitation to enroll in this prospective study. As in our previous studies, we excluded participants only being treated for conditions such as osteoarthritis, fibromyalgia, mechanical back pain, gout, or pseudogout(2, 9, 11-14).

### Prospective study on sequelae of COVID-19

We invited all SARDs participants with confirmed COVID-19 to participate in a prospective, longitudinal study, COVID-19 and Rheumatic Diseases (RheumCARD), to collect surveys and biospecimens. Potential participants (excluding those who died from COVID-19) were contacted either via secure online participant portal or US mail to determine willingness to participate in the study. The initial invitations were sent on March 11, 2021. The recruitment material and surveys were made available in Spanish starting October 12, 2021. Invitations to RheumCARD were sent out every 2-4 weeks on a rolling basis as newly infected SARD participants with confirmed COVID-19 were identified. This study analyzed the RheumCARD baseline survey that collected data related to demographics, baseline clinical characteristics, COVID-19 symptoms and disease course, COVID-19 vaccination status, and SARD medications before and after COVID-19. The study was approved by the Mass General Brigham Institutional Review Board.

### Demographics, SARD characteristics, comorbidities, and COVID-19 course

Demographics assessed in the survey included age, sex, and race/ethnicity. Data on smoking status at the time of survey (never, past, current) were collected. Participant-reported comorbidities included obesity, hypertension, asthma, obstructive sleep apnea, coronary artery disease, diabetes, heart failure, chronic kidney disease, chronic obstructive pulmonary disease, peptic ulcer disease, pulmonary hypertension, solid tumor, stroke, acquired immunodeficiency syndrome, cirrhosis, dementia, hemiplegia, interstitial lung disease/pulmonary fibrosis, leukemia, lymphoma, and peripheral vascular disease. The comorbidity count was derived as the sum of these comorbidities that were assessed in the survey.

Dates of positive SARS-CoV-2 test and COVID-19 symptom onset were collected. Symptoms assessed in the survey included: fever, sore throat, new cough, nasal congestion/rhinorrhea, dyspnea, chest pain, rash, myalgias, fatigue/malaise, headache, nausea/vomiting, diarrhea, anosmia, dysgeusia, and joint pain. The COVID-19 initial symptom count was defined as the sum of these symptoms assessed at time of survey. Acute COVID-19 course, including treatment (glucocorticoids, remdesivir, neutralizing monoclonal antibodies to SARS-CoV-2, or clinical trial enrollment), hospitalization, supplemental oxygen, and mechanical ventilation were obtained by survey. Time to COVID-19 symptom resolution, type of persistent symptoms (same list as assessed from initial symptoms), and vaccination status (before infection, after infection, declined, and planning to receive) at time of survey were also assessed by self-report.

We also assessed for baseline glucocorticoid and DMARD use at the time of COVID-19 diagnosis. Immunomodulator medications included conventional synthetic (cs) DMARDs (methotrexate, hydroxychloroquine, sulfasalazine, azathioprine, leflunomide, mycophenolate mofetil), tumor necrosis factor inhibitors (TNFi: adalimumab, certolizumab pegol, etanercept, golimumab, infliximab), rituximab, ustekinumab, tocilizumab, abatacept, secukinumab, belimumab, apremilast, upadacitinib, tofacitinib, and intravenous immunoglobulin. Participants also reported the following characteristics about changes in DMARDs around COVID-19 diagnosis for each medication: temporarily stopped, continued, increased dose, decreased dose, and decreased frequency.

### SARD activity/flare, pain, fatigue, functional status, and respiratory quality of life

Participants were asked whether their underlying SARD flared after COVID-19. For those who flared, the timing after COVID-19 was also obtained (<1, 1-4, 4-12, or >12 weeks). A global assessment of rheumatic disease activity was also obtained where 0 represented “very poor” and 10 represented “very well” disease control. Participants rated their disease activity just prior to COVID-19 onset as well as at the time of survey.

The survey included several validated survey instruments to collect information on SARD activity, pain, fatigue, quality of life, and respiratory symptoms. The Routine Assessment of Participant Index Data 3 (RAPID3) was used to assess SARD activity (15). A score was calculated to indicate level of disease activity, as previously described (0 [remission], 0.3-1.0 [near remission], 1.3-2.0 [low severity], 2.3-4.0 [moderate severity], 4.3-10.0 [high severity]) (16). The visual analog and rank value scales from the Short-Form McGill Pain Questionnaire (SF-MPQ) were utilized to assess respondents’ pain(17). Fatigue was quantified using the Fatigue Symptom Inventory (FSI)(18). Participants completed the modified Health Assessment Questionnaire (mHAQ) to assess functional status(19). Scores were divided into categories based on severity of impairment (<1.3 mild, 1.3-1.8 moderate, >1.8 severe), as previously described(20). Finally, the St. George’s Respiratory Questionnaire (SGRQ) was used to measure respiratory quality of life(21). We calculated the global score as well as individual SGRQ components for symptoms, activity, and impact, as previously described(21).

### Prolonged symptom duration definition

As in general population studies, prolonged symptom duration (or PASC) was defined either as those suffering from COVID-19 symptoms persistently for 28 or more days, or those whose symptoms initially resolved (or were not initially present) but then recurred/developed 28 or more days after COVID-19 onset, as defined by the US Centers for Disease Control and Prevention (CDC)(22, 23).

### Statistical analysis

We used descriptive statistics to summarize variables. We analyzed the entire study sample and also stratified by presence/absence of prolonged COVID-19 symptom duration of at least 28 days (i.e., PASC). We compared characteristics of acute COVID-19 course, persistent COVID-19 symptoms/duration, SARD flare/activity, RAPID-3 scores, SF-MPQ scores, FSI scores, and SGRQ scores. For categorical variables, we obtained p values using chi-square or Fisher’s exact (for variables with low cell sizes) tests. Wilcoxon rank-sum test was used to obtain p values for continuous variables. The Wilcoxon signed rank test was used to assess for a difference in participant-reported SARD control, comparing participant global assessment before COVID-19 onset to the time of survey.

Unadjusted logistic regression was performed to estimate odds ratios (ORs) and 95% confidence intervals (CIs) for prolonged symptom duration by baseline factors. We considered demographic variables and any baseline (i.e., at COVID-19 onset) variables with p values <0.1 on univariate testing. We considered the initial COVID-19 symptom count rather than specific symptoms since several were associated in unadjusted analyses. Multivariable models additionally adjusted for age, sex, race, smoking, and comorbidity count. All analyses were performed using SAS v.9.4 (Cary, NC). We considered two-sided p<0.05 as statistically significant.

## RESULTS

### Demographics and clinical characteristics

As of January 5, 2022, 670 surveys were sent to SARD participants confirmed to have survived COVID-19. We analyzed survey responses from 174 participants (response rate 26%) who completed the survey (**Figure 1**). Female sex and White race were associated with increased likelihood to respond to the survey (**Supplemental Table 1**). A smaller proportion of responders were hospitalized compared to non-responders, but this was not statistically significant (18% vs. 25%, p=0.085).

**Figure 1.**
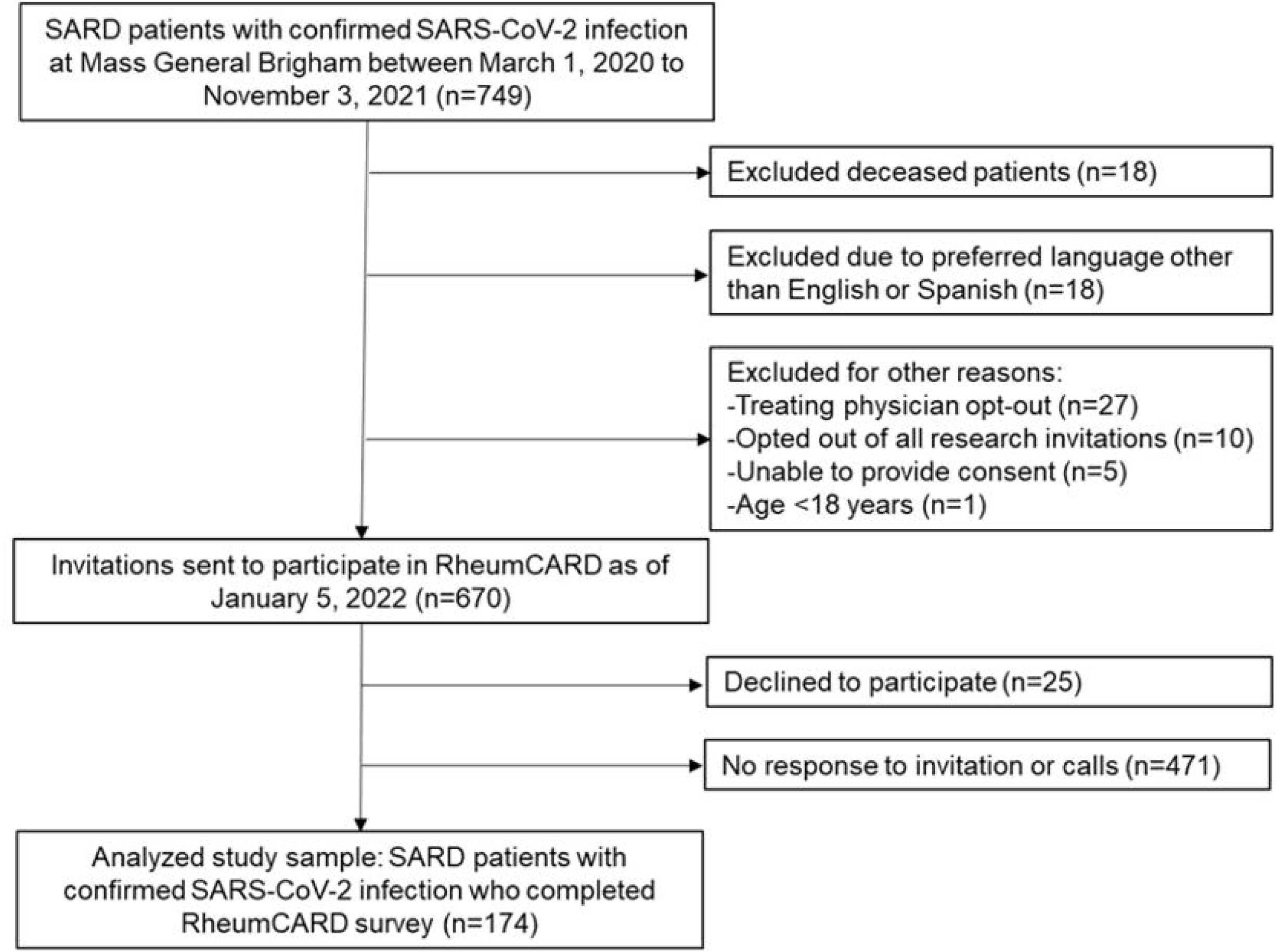
Flow chart describing recruitment and analyzed study sample. RheumCARD, COVID-19 in Autoimmune Rheumatic Disease; SARD, systemic autoimmune rheumatic disease; SARS-CoV-2, severe acute respiratory syndrome coronavirus 2.

Mean age and standard deviation (SD) for the sample was 52 ± 16 years (**Table 1**). Most respondents were female (81%), White (80%), and had never smoked (72%). The most common SARD was RA (40%), followed by SLE (14%) and PsA (12%). The most common comorbidities were obesity (22%), hypertension (20%), and asthma (14%).

**Table 1.** Demographics and clinical characteristics of COVID-19 survivors with SARDs who provided survey data (n=174).

|  |  |
| --- | --- |
| Age at time of survey in years, mean ± SD | 52 ± 16 |
| Female, n (%) | 140 (81) |
| Race, n (%) |  |
| White | 139 (80) |
| Black | 17 (10) |
| Asian | 9 (5) |
| Other | 7 (4) |
| Hispanic or Latinx ethnicity, n (%) | 18 (10) |
| Smoking status*, n (%) |  |
| Never | 125 (72) |
| Past | 47 (27) |
| Current | 1 (<1) |
| Type of SARD**, n (%) |  |
| Rheumatoid arthritis | 69 (40) |
| Systemic lupus erythematosus | 25 (14) |
| Psoriatic arthritis | 21 (12) |
| Sjögren's syndrome | 10 (6) |
| Axial spondyloarthritis | 9 (5) |
| Polymyalgia rheumatica | 8 (5) |
| ANCA-associated vasculitis | 6 (3) |
| Mixed connective tissue disease | 5 (3) |
| Inflammatory myositis | 3 (2) |
| Juvenile idiopathic arthritis | 3 (2) |
| Systemic sclerosis | 2 (1) |
| Sarcoidosis | 2 (1) |
| Baseline glucocorticoid use at COVID-19 onset, n (%) | 29 (18) |
| Dose in daily mg of prednisone equivalents, median [IQR] | 9.5 [5, 13.5] |
| Comorbidities***, n (%) |  |
| Obesity | 39 (22) |
| Hypertension | 35 (20) |
| Asthma | 25 (14) |
| Obstructive sleep apnea | 16 (9) |
| Coronary artery disease | 12 (7) |
| Diabetes | 11 (6) |
| Heart failure | 5 (3) |
| Chronic kidney disease | 5 (3) |
| Chronic obstructive pulmonary disease | 3 (2) |
| Interstitial lung disease/pulmonary fibrosis | 3 (2) |
| Solid tumor | 3 (2) |
| Comorbidity count#, median [IQR] | 1 [0, 2] |

ANCA, anti-neutrophil cytoplasmic antibody; COVID-19, coronavirus disease 2019; IQR, interquartile range; SARD, systemic autoimmune rheumatic disease; SD, standard deviation.
\*Smoking status was missing for 1 participant.
\*\*Other SARDs included IgG4-related disease, undifferentiated connective tissue disease, giant cell arteritis (each n=1).
\*\*\*Other comorbidities included peptic ulcer disease, pulmonary hypertension, stroke, and dementia (each n=2).
#Defined as the sum of comorbidities assessed in the survey.

### Acute COVID-19 course

The median time from COVID-19 onset to survey response was 230 [165, 360] days. (**Table 2**). The most common acute symptoms were fatigue/malaise (72%), fever (60%), and headache (60%). Significantly more participants with vs. without prolonged symptoms experienced fatigue (87% vs. 59%, p<0.001), fever (73% vs. 49%, p=0.001), headache (76% vs. 47%, p<0.001), myalgias (74% vs. 42%, p<0.001), anosmia (56% vs. 33%, p=0.002), dysgeusia (60% vs. 23%, p<0.001), and dyspnea (44% vs. 21%, p=0.001). Those with symptoms ≥28 days exhibited a significantly higher median initial symptom count (7 [6, 9] vs. 4 [2, 7], p<0.001).

**Table 2.**
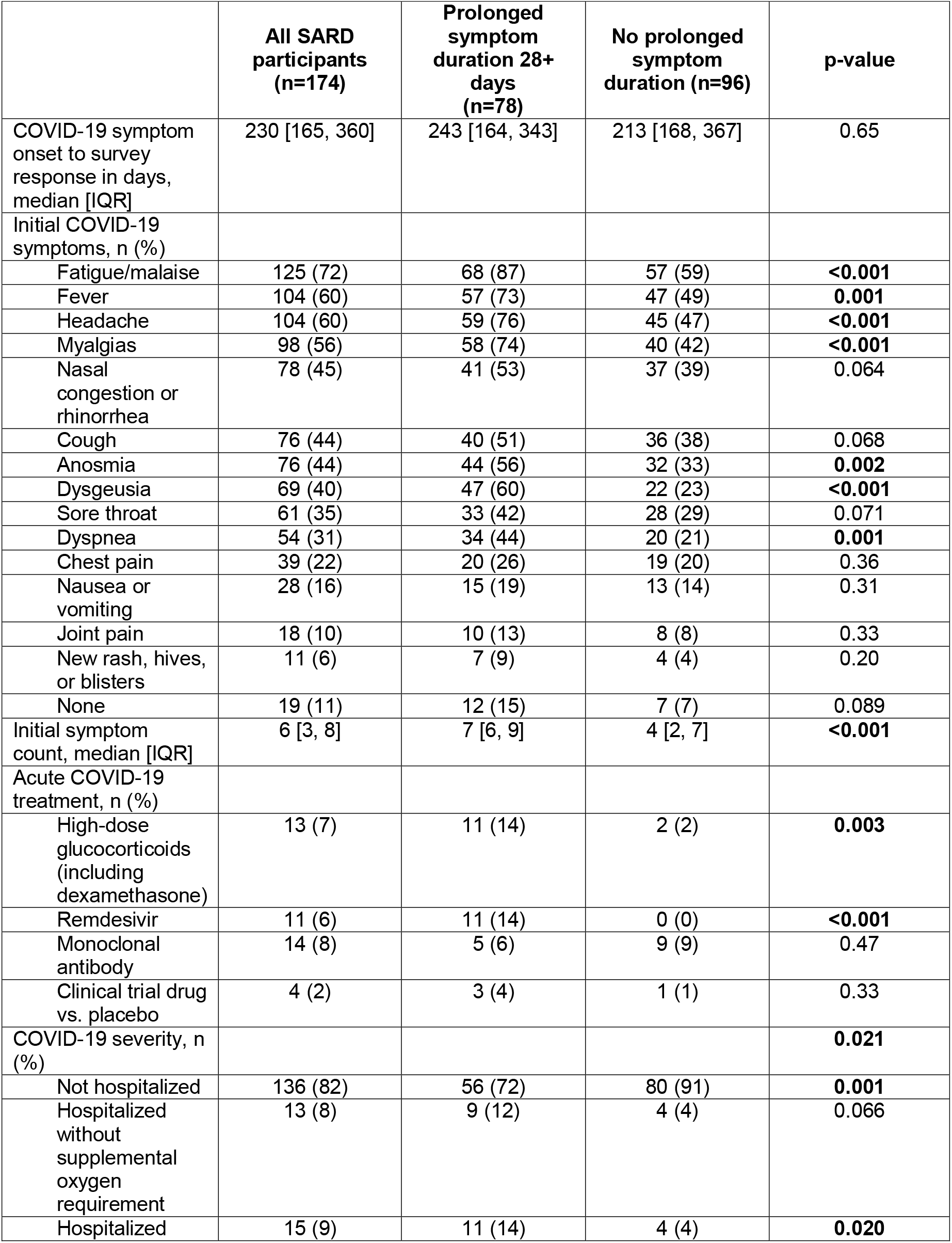
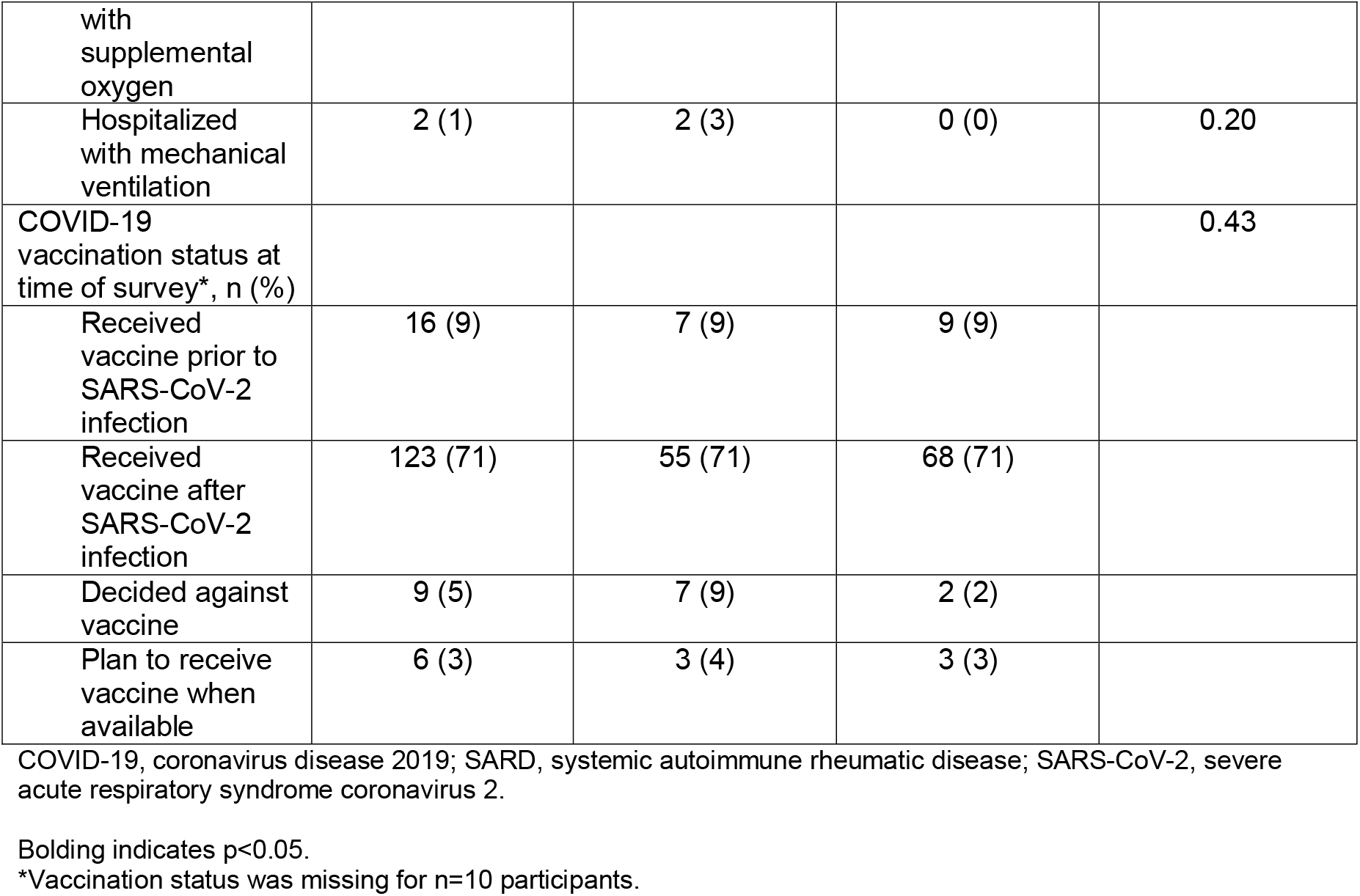
Acute COVID-19 course and vaccination status for the entire study sample, and for those with and without prolonged symptom duration.

The group with prolonged symptoms had higher proportions of hospitalization (28% vs. 9%, p=0.001) as well as hospitalization with supplemental oxygen (14% vs. 4%, p=0.020). Those with prolonged symptom duration were also more likely to have received high-dose glucocorticoids (14% vs. 2%, p=0.003) and remdesivir (14% vs. 0%, p<0.001) to treat COVID-19. Most participants received COVID-19 vaccination after infection (79% vs. 80%, p=0.43). Sixteen participants reported breakthrough infection occurring after COVID-19 vaccination.

### Immunosuppressive medication use and disruption after COVID-19

Eighteen percent of participants reported taking glucocorticoids at COVID-19 onset (median [IQR] daily prednisone-equivalents of 9.5 mg [5, 13.5]. DMARDs were prescribed to most respondents at time of survey (153 DMARDs prescribed among 127 unique participants; **Figure 2**). Over half (65/127, 51%) of patients on any DMARDs made any change, mostly temporarily stopping (49/127, 39%), followed by stretching the dosing interval (10/127, 8%), decreasing the dose (5/127, 4%), and starting a new DMARD (2/127, 2%). When analyzing individual medications, most had high rates of disruption. For example, TNFi was disrupted in 27/35 (77%) and methotrexate in 24/32 (75%). However, hydroxychloroquine was disrupted in only 6/23 (23%) and rituximab in only 6/13 (46%). After excluding hydroxychloroquine, 30/42 (71%) of csDMARDs were disrupted. After excluding rituximab, 38/52 (73%) of bDMARDs were disrupted.

**Figure 2.**
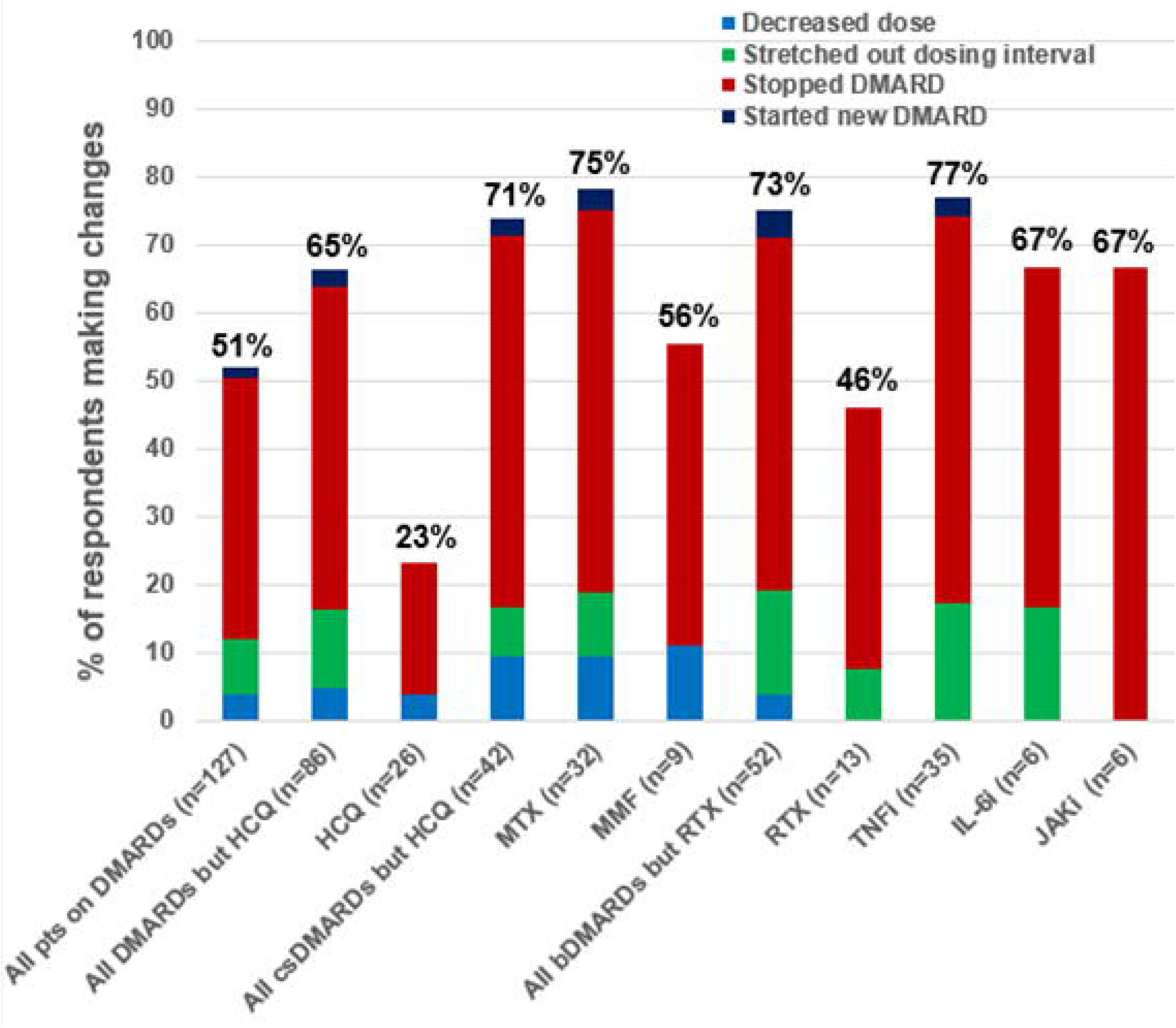
Frequencies of baseline DMARD use and proportions with any disruption at COVID-19 onset. bDMARDs, biologic disease-modifying antirheumatic drugs; csDMARDs, conventional synthetic disease-modifying antirheumatic drugs DMARDs, disease-modifying antirheumatic drugs; HCQ, hydroxychloroquine; IL-6i, interleukin-6 inhibitors; JAKi, Janus kinase inhibitors; MMF, mycophenolate mofetil; MTX, methotrexate; RTX, rituximab; TNFi, tumor necrosis factor inhibitors.

### Lingering COVID-19 symptoms and SARD activity after SARS-CoV-2 infection

The median symptom duration in all participants was 14 days (IQR 9, 29; **Table 3**). There were 78 participants (45%) that met the CDC definition for prolonged symptom duration (28 or more days after COVID-19 onset). Those with prolonged symptom duration had a median of 46 [30, 65] days to COVID-19 symptom duration compared to 11 [7, 14] for those without prolonged symptom duration (p<0.0001). The most common persistent symptoms among 54 participants with prolonged duration were fatigue (27%), anosmia (18%), dysgeusia (17%), dyspnea (12%), and nasal congestion (12%).

**Table 3.**
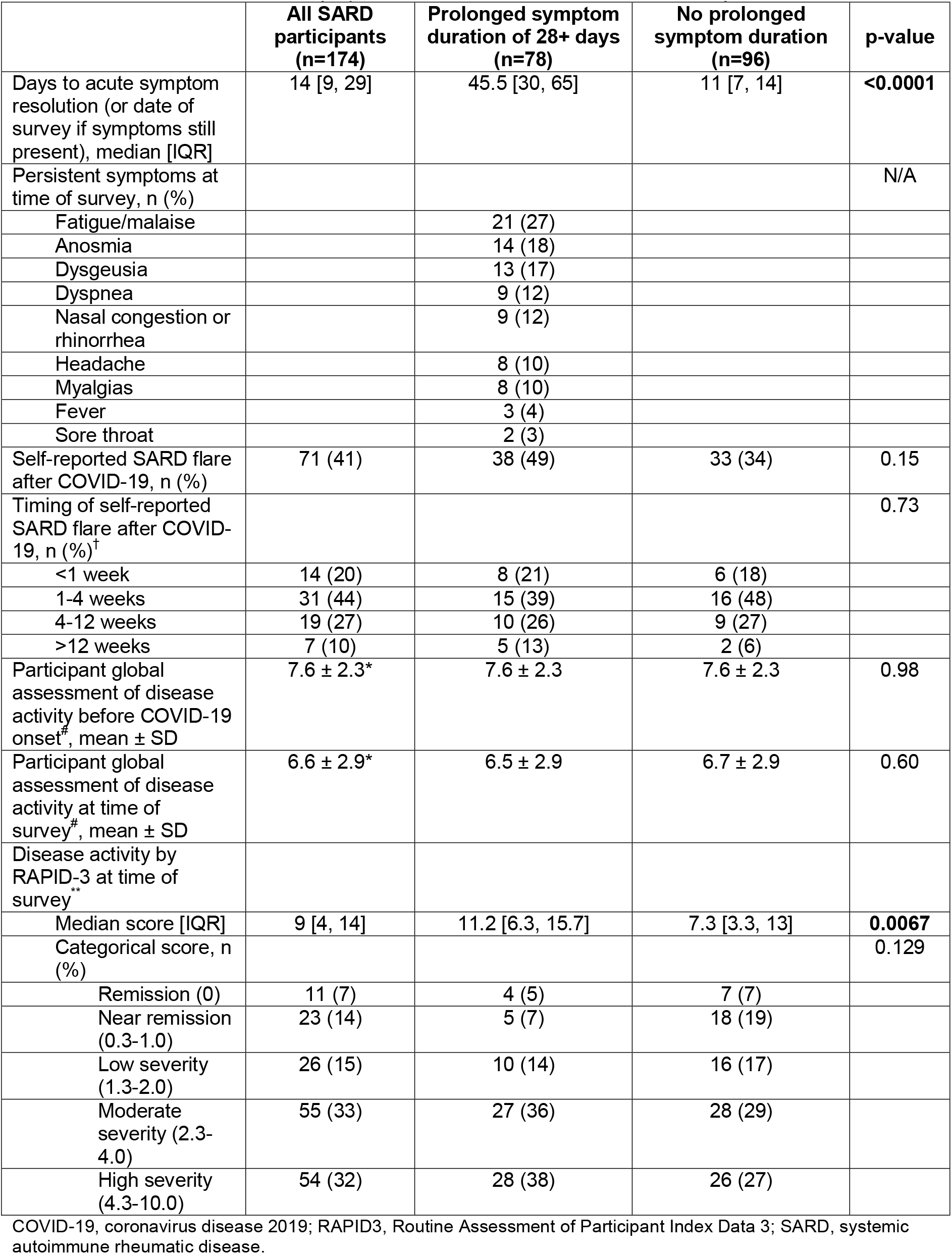
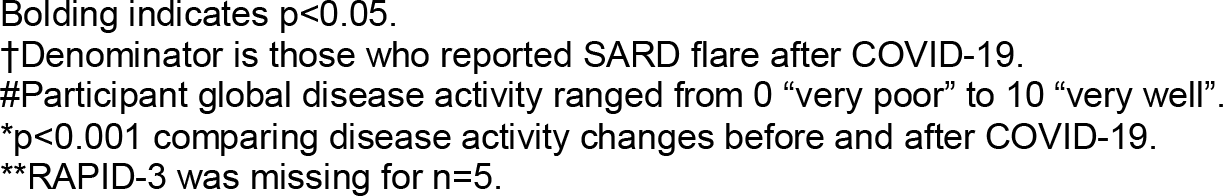
Persistent COVID-19 symptoms/duration and SARD flares/activity after COVID-19.

|  | **All SARD participants**<br>**(n=174)** | **Prolonged symptom duration of 28+ days**<br>**(n=78)** | **No prolonged symptom duration**<br>**(n=96)** | **p-value** |
| --- | --- | --- | --- | --- |
| Days to acute symptom resolution (or date of survey if symptoms still present), median [IQR] | 14 [9, 29] | 45.5 [30, 65] | 11 [7, 14] | **<0.0001** |
| Persistent symptoms at time of survey, n (%) |  |  |  | N/A |
| Fatigue/malaise |  | 21 (27) |  |  |
| Anosmia |  | 14 (18) |  |  |
| Dysgeusia |  | 13 (17) |  |  |
| Dyspnea |  | 9 (12) |  |  |
| Nasal congestion or rhinorrhea |  | 9 (12) |  |  |
| Headache |  | 8 (10) |  |  |
| Myalgias |  | 8 (10) |  |  |
| Fever |  | 3 (4) |  |  |
| Sore throat |  | 2 (3) |  |  |
| Self-reported SARD flare after COVID-19, n (%) | 71 (41) | 38 (49) | 33 (34) | 0.15 |
| Timing of self-reported SARD flare after COVID-19, n (%)<sup>†</sup> |  |  |  | 0.73 |
| <1 week | 14 (20) | 8 (21) | 6 (18) |  |
| 1-4 weeks | 31 (44) | 15 (39) | 16 (48) |  |
| 4-12 weeks | 19 (27) | 10 (26) | 9 (27) |  |
| >12 weeks | 7 (10) | 5 (13) | 2 (6) |  |
| Participant global assessment of disease activity before COVID-19 onset<sup>#</sup>, mean ± SD | 7.6 ± 2.3* | 7.6 ± 2.3 | 7.6 ± 2.3 | 0.98 |
| Participant global assessment of disease activity at time of survey<sup>#</sup>, mean ± SD | 6.6 ± 2.9* | 6.5 ± 2.9 | 6.7 ± 2.9 | 0.60 |
| Disease activity by RAPID-3 at time of survey<sup>**</sup> |  |  |  |  |
| Median score [IQR] | 9 [4, 14] | 11.2 [6.3, 15.7] | 7.3 [3.3, 13] | **0.0067** |
| Categorical score, n (%) |  |  |  | 0.129 |
| Remission (0) | 11 (7) | 4 (5) | 7 (7) |  |
| Near remission (0.3-1.0) | 23 (14) | 5 (7) | 18 (19) |  |
| Low severity (1.3-2.0) | 26 (15) | 10 (14) | 16 (17) |  |
| Moderate severity (2.3-4.0) | 55 (33) | 27 (36) | 28 (29) |  |
| High severity (4.3-10.0) | 54 (32) | 28 (38) | 26 (27) |  |

COVID-19, coronavirus disease 2019; RAPID3, Routine Assessment of Participant Index Data 3; SARD, systemic autoimmune rheumatic disease.
Bolding indicates $p < 0.05$ .
†Denominator is those who reported SARD flare after COVID-19.

Participants experienced significantly worse rheumatic disease control after (mean ± SD global assessment of disease activity 6.6 ± 2.9) compared to before COVID-19 (7.6 ± 2.3) (p<0.001; higher scores indicating better self-reported disease control). Forty-one percent of respondents reported SARD flare after COVID-19, with most occurring 1-4 weeks after diagnosis (44%). The median RAPID3 score (higher scores indicating higher disease activity) for the group with prolonged symptom duration was 11.2 [6.3, 15.7] compared to 7.3 [3.3, 13] for the group without prolonged symptom duration (p=0.0067).

### Predictors of prolonged COVID-19 symptom duration

In the unadjusted logistic regression model, hospitalization and symptom count had ORs of 3.93 (95% confidence interval [CI] 1.63-9.46) and 1.33 (per symptom, 95%CI 1.19, 1.49) for prolonged symptom duration, respectively. In the multivariable model, hospitalization for COVID-19 had an OR of 3.54 (95%CI 1.27, 9.87) for symptom duration of 28 or more days, while symptom count had an OR of 1.35 (per symptom, 95%CI 1.18, 1.55) after adjusting for age, sex, race, smoking, and comorbidity count.

### Pain, fatigue, functional status, and respiratory quality of life after COVID-19

Participants with prolonged symptom duration recorded a significantly higher median SF-MPQ score (indicating more pain) compared to the other group (2 [1,2] vs. 1 [0,2], p=0.0008) (**Table 4**). The median FSI was significantly higher among participants with prolonged COVID-19 symptom duration (66 [31, 91.5] vs. 43 [26, 76], p=0.031; higher scores indicating more fatigue). There was no association of prolonged symptom duration with mHAQ (median 0.25 [0, 0.75] vs. 0.125 [0, 0.0375], p=0.11). The median global SGRQ score was 14.7 (IQR 4.3, 29.3; higher scores indicating worse respiratory quality of life). Those with prolonged symptom duration had a median score of 16.1 compared to 10.1 for the other participants, but these values were not statistically different (p=0.49). There were also no statistical differences for those with and without prolonged symptom duration in SGRQ subscales that included symptoms, activity, and impact scores.

**Table 4.** Pain, fatigue, functional status, and respiratory quality of life of COVID-19 survivors with SARDs at time of survey.

|  | **All SARD participants**<br>**(n=174)** | **Prolonged symptom duration of 28+ days**<br>**(n=78)** | **No prolonged symptom duration**<br>**(n=96)** | **p-value** |
| --- | --- | --- | --- | --- |
| **Pain (SF-MPQ)\*** |  |  |  |  |
| Median score [IQR] | 2 [1, 2] | 2 [1, 2] | 1 [0, 2] | **0.0008** |
| Pain rating index ordinal categories, n (%) |  |  |  | **0.011** |
| No pain | 31 (19) | 8 (11) | 23 (25) |  |
| Mild pain | 46 (29) | 15 (21) | 31 (34) |  |
| Discomforting pain | 64 (40) | 35 (50) | 29 (32) |  |
| Distressing pain | 14 (9) | 7 (10) | 7 (8) |  |
| Horrible pain | 4 (2) | 4 (6) | 0 (0) |  |
| Excruciating pain | 2 (1) | 1 (1) | 1 (1) |  |
| **Fatigue (FSI)** |  |  |  |  |
| Median score [IQR] | 53 [27, 84] | 66 [31, 91.5] | 43 [26, 76] | **0.031** |
| Categorical score (quartiles), n (%) |  |  |  | 0.094 |
| Q1 ($\leq 26$; least fatigued) | 51 (29) | 23 (29) | 28 (29) | |
| Q2 (27-54) | 47 (27) | 15 (19) | 32 (33) |  |
| Q3 (55-85) | 39 (22) | 18 (23) | 21 (22) |  |
| Q4 ($\geq 86$; most fatigued) | 37 (21) | 22 (28) | 15 (16) | |
| **Functional status (mHAQ)** |  |  |  |  |
| Median score [IQR] | 0.125 [0, 0.375] | 0.25 [0, 0.75] | 0.125 [0, 0.375] | 0.11 |
| Categorical score, n (%) |  |  |  | 0.27 |
| Normal ($<0.3$) | 109 (63) | 44 (56) | 65 (68) | |
| Mild impairment (0.3-1.3) | 62 (36) | 32 (41) | 30 (31) |  |
| Moderate impairment (1.3-1.8) | 3 (2) | 2 (3) | 1 (1) |  |
| Severe impairment ($>1.8$) | 0 (0) | 0 (0) | 0 (0) | |
| **Respiratory quality of life (SGRQ)\*\*** |  |  |  |  |
| Global score, median [IQR] | 14.7 [4.3, 29.3] | 16.1 [4.4, 36.1] | 10.1 [4.1, 25.6] | 0.49 |
| Symptoms score, median [IQR] | 13.7 [0, 31.8] | 14.2 [0, 33.2] | 13.6 [0, 29.3] | 0.79 |
| Activity score, median [IQR] | 29.6 [0, 47.7] | 35.8 [0, 57.2] | 25.2 [0, 42.4] | 0.19 |
| Impact score, median [IQR] | 4.6 [0, 15.2] | 4.8 [0, 16.1] | 4.1 [0, 13.1] | 0.54 |

SF-MPQ, Short-form McGill Pain Questionnaire; FSI, Fatigue Symptom Inventory; mHAQ, modified Health Assessment Questionnaire; SGRQ, Saint George's Respiratory Questionnaire.
Bolding indicates $p < 0.05$ .
\*SF-MPQ was missing for 13 (7%) participants.
\*\*SGRQ was missing for 58 (33%) participants.

## DISCUSSION

In this prospective study, we describe a potentially substantial impact of post-acute sequelae after acute COVID-19 for patients with SARDs. A high proportion of DMARDs were disrupted at COVID-19 onset, ranging between 56-77%. Two out of five participants reported a flare of their underlying SARD following COVID-19. There was a statistically significant worsening in self-reported disease control even months after COVID-19 onset. A substantial proportion (45%) reported prolonged duration of COVID-19 symptoms (at least 28 days). This may indicate that participants with SARDs may be particularly vulnerable to persistent worsening quality of life (more pain and fatigue), and potentially morbidity, even beyond the acute period. Collectively, our findings suggest a large health impact of COVID-19 on participants with SARDs, even after the acute infection.

There has been intense interest in understanding whether SARDs participants are at increased risk for severe acute COVID-19 outcomes, such as hospitalization, mechanical ventilation, and death(1-4). Indeed, SARDs participants do seem to have worse acute COVID-19 outcomes than the general population, perhaps related to altered underlying immunity, propensity for hyperinflammation, immunosuppression, organ damage from underlying SARD, and comorbidities(24). Many of the same factors that place SARDs at risk of poor acute outcomes could place them at risk of having a protracted COVID-19 course even if they are no longer at risk for COVID-19-related hospitalization or death. However, there has been relatively less attention to post-acute sequelae of COVID-19 among SARDs despite high reported rates of rheumatic and musculoskeletal manifestations in the general population(25-27). Since PASC and SARDs share many potential pathogenic features, such as systemic inflammation, fibrosis, autoimmunity, and hypercoagulability, uncovering the mechanisms of PASC in SARDs may have public health implications even for the general population(28).

An early study surveyed 105 rheumatology participants in Spain who had been hospitalized with COVID-19 and classified them as having SARDs (n=54) or non-autoimmune rheumatic conditions (n=51, such as osteoarthritis)(29). Similar to our study, they found that a high proportion of subjects had lingering symptoms (some lasting for over 90 days) that included dyspnea, fatigue, chest pain, and cough. Several participants had new clinically apparent lung damage (10%), and 6% had a new oxygen requirement that persisted as an outpatient. Our study extends these findings by including non-hospitalized SARDs participants, which is important since most COVID-19 cases are managed as an outpatient. We also identified dyspnea as a common persistent symptom, emphasizing the lung as a potential site for damage vulnerable in both acute COVID-19 and PASC(2, 12, 30). Future studies are needed to investigate whether SARD participants may be more prone to residual lung damage after COVID-19, perhaps related to pre-existing subclinical lung damage or underlying propensity for pulmonary fibrosis and interstitial lung disease in SARDs.

A more recent study surveyed participants seen at an academic rheumatology center in New York City and compared “long haul” COVID-19 (symptoms for 3+ months, n=142) to non-long haul COVID-19 (<1 month symptoms, n=112)(31). COVID-19 was identified by self-report of a positive test or suspected by a physician. About 60% of these respondents had a SARD diagnosis. Similar to our findings, they found that higher burden of initial COVID-19 symptoms was associated with long-hauler status. They also found worse quality of life on participant-Reported Outcomes Measurement Information System measures that included pain (similar to our study), fatigue, anxiety, and depression. Fatigue was commonly reported in our study as well and though this may reflect contributions from underlying SARD, it is notable that our survey asks participants to specifically identify symptoms of their acute COVID-19 infection and identify those which persist. Our study also found a high proportion of participants with SARDs experienced prolonged COVID-19 symptom duration. Many participants in our study reported fatigue of which both COVID-19 and underlying SARD may have contributed. However, many also reported dysgeusia and anosmia, likely more specifically related to COVID-19. Additionally, the patient’s global assessment of rheumatic disease control was similar between those with and without prolonged symptom duration, suggesting other contributing factors driving these observed differences in symptoms following acute COVID-19.

To our knowledge, ours is the first to describe DMARD disruptions and SARD flare/activity after acute COVID-19. We found that over half of participants either temporarily discontinued or decreased the dose/frequency of DMARDs at acute COVID-19 diagnosis. We also found that nearly half of participants reported a flare of underlying SARD in the weeks following acute infection and participants had a statistical worsening of self-reported global assessment of disease activity after COVID-19 at the time of survey. Medication disruption may have contributed to flare and increased disease activity that persisted through to the time of survey completion. Prior studies have focused mainly on DMARD changes early in the pandemic related to concerns about drug supply or immunosuppression(32-34). Other studies have focused on temporary medication changes after COVID-19 vaccination and have found little evidence of increased risk of flare(35-38). Compared to those studies, participants in our study may have had longer disruption of DMARDs while acutely ill, particularly those who required hospitalization, which may have increased their risk of flare. Additionally, it is possible that participants in our study may have had immune system activation as a reaction to SARS-CoV-2 infection that could have contributed to increased activity in addition to DMARD disruption. Indeed, we previously found that participants with SARDs hospitalized for COVID-19 were more prone to hyperinflammation(12). Finally, many participants in the general population report widespread pain and arthralgias after COVID-19, which may have also influenced the high rate of self-reported SARD flare(25-27). We found that participants with persistent symptom duration had higher levels of pain than those with resolved symptoms. Future studies are needed to include comparators with SARDs but no history of COVID-19 as well as general population comparators with COVID-19 to better understand these complex relationships between COVID-19 infection, underlying SARD flare, and worsened pain or fatigue.

Strengths of our study include its prospective nature, inclusion of participants across the spectrum of acute COVID-19 severity, and the systematic approach towards identifying and confirming both COVID-19 and SARD. This study focused on participant-reported outcome measures as one of the first comprehensive assessments of post-acute sequelae of COVID-19 in SARDs participants recruited from several rheumatology centers within a large health care system. We assessed many validated survey instruments including the RAPID-3, SF-MPQ, FSI, mHAQ, and SGRQ to capture the myriad potential effects of COVID-19 infection on outcomes particularly relevant to SARDs participants. Ours is the first to describe DMARD disruption, underlying SARD flare/activity, and prolonged COVID-19 symptom duration. We focused on all survivors of COVID-19, not only those who required hospitalization.

Our study does have limitations. The cross-sectional nature allowed for the surveying of respondents at only one point in time post-COVID-19, possibly introducing recall bias, particularly related to assessment of disease activity and symptoms at acute COVID-19 onset. The survey response rate was 26%, similar to other survey studies, but there were some factors that may have response rate and could affect generalizability. We were also only able to assess survivors of COVID-19. Some participants who were very ill during their acute COVID-19 course may have been less likely to respond to the survey while recuperating. We found that hospitalization was significantly associated with prolonged symptom duration. Future studies may focus on this vulnerable population at high risk for protracted COVID-19 course. Our sample was heterogeneous, composed of participants with a variety of different SARDs. Future analyses in this study will focus on specific SARDs such as RA once the sample size is sufficient. Disease activity and flare were obtained by participant report rather than in-person physical examination. Since many SARD types were included, a single validated assessment would have been difficult to measure. However, the RAPID3 is commonly used as a survey instrument for all SARD participants to measure participant-reported disease activity and pain. It is possible that a small proportion of participants may have had persistent viral infection, as has been described for SARDs with COVID-19 on B cell depleting therapy(39). We were unable to investigate autoantibody expansion and increased systemic inflammation using only survey data. Future directions of RheumCARD will address many of these limitations. These include the addition of a comparator group with identical assessments but no history of COVID-19, biospecimen sample collection (including serum, plasma, peripheral blood mononuclear cells, and nasopharyngeal swabs), survey assessments at multiple points in time, and initial enrollment early in the COVID-19 course.

In summary, we found that COVID-19 survivors with SARDs commonly experienced DMARD disruptions, SARD flares, and high self-reported disease activity. Additionally, two out of five SARD participants experienced prolonged duration of COVID-19 symptoms lasting 28 or more days that included fatigue, dysgeusia, anosmia, and dyspnea and higher levels of pain and fatigue than those without prolonged symptom duration. Even after resolution of acute COVID-19, many participants with SARDs may continue to experience effects related to COVID-19 and may also significantly impact their underlying diseases. The results of this study and those that follow will serve to inform the approach towards participants with SARDs who have experienced COVID-19, both in management of their underlying rheumatic diseases as well as their long-term COVID-19 symptoms.

## Data Availability

All data produced in the present study are available upon reasonable request to the authors.

