## Supplemental Tables for "DMARD disruption, disease flare, and prolonged symptom duration after acute COVID-19 among participants with rheumatic disease: A prospective study"

| **Supplemental Table 1.** Comparison of survey respondents to invitees who did not respond to the survey. | | | |
| --- | --- | --- | --- |
|  | **Responded to survey (n=174)** | **Invited but did not respond to survey (n=492)** | **p-value** |
| Mean age, years ± SD | 52 ± 16 | 55 ± 16 | 0.078 |
| Female sex, n (%) | 140 (81) | 360 (73) | **0.042** |
| Race |  |  |  |
| White, n (%) | 139 (80) | 330 (67) | **0.0015** |
| Black, n (%) | 17 (10) | 65 (13) | 0.24 |
| Asian, n (%) | 9 (5) | 12 (2) | 0.082 |
| Other, n (%) | 7 (4) | 86 (17) | **<0.0001** |
| Hospitalized for COVID-19, n (%) | 30 (18) | 103 (25) | 0.085 |
| Bolding indicates p<0.05. | | | |

| **Supplemental Table 2.** Unadjusted and multivariable odds ratios for prolonged COVID-19 symptom duration of 28+ days in COVID-19 survivors with SARDs (n=174). | | |
| --- | --- | --- |
| **Covariates** | **Unadjusted OR (95% CI) for prolonged symptom duration of 28+ days** | **Multivariable OR (95% CI) for prolonged symptom duration of 28+ days** |
| Hospitalized (vs. not) | **3.93 (1.63, 9.46)** | **3.54 (1.27, 9.87)** |
| Initial symptom count at COVID-19 onset (per symptom) | **1.33 (1.19, 1.49)** | **1.35 (1.18, 1.55)** |
| Age (per year) | 1.00 (0.98, 1.02) | 1.01 (0.98, 1.04) |
| Female (vs. male) | 1.56 (0.71, 3.42) | 1.00 (0.38, 2.61) |
| White (vs. non-White) | 1.49 (0.69, 3.19) | 1.42 (0.29, 6.96) |
| Black (vs. non-Black) | 0.64 (0.23, 1.83) | 0.53 (0.08, 3.47) |
| Asian (vs. non-Asian) | 0.14 (0.02, 1.17) | 0.23 (0.01, 4.37) |
| Smoking (ever vs. never) | 1.87 (0.95, 3.66) | 1.49 (0.63, 3.55) |
| Comorbidity count (per comorbidity) | 1.04 (0.83, 1.32) | 0.98 (0.73, 1.31) |
| CI confidence interval; COVID-19; coronavirus disease 2019; OR, odds ratio.  Bolding indicates p<0.05. | | |
